# An Ebola virus disease model with fear and environmental transmission dynamics

**DOI:** 10.1101/2021.01.05.20249082

**Authors:** M.L. Juga, F. Nyabadza, F. Chirove

## Abstract

Recent Ebola virus disease (EVD) outbreaks have been limited not only to the interactions between humans but also to the complex interplay of the environment, human and socio-economic factors. Changes in human behaviour as a result of fear can also affect disease transmission dynamics. In this paper, a compartmental model is used to study the dynamics of EVD incorporating fear and environmental transmission. We formulate a fear dependent contact rate function to measure the rate of person to person, as well as pathogen to person transmissions. The epidemic threshold and the model equilibria are determined and, their stabilities are analysed. The model is validated by fitting it to data from the 2019 and 2020 EVD outbreaks in the Democratic Republic of Congo. Our results suggest that the fear of death from EVD may reduce the transmission and aid the control of the disease, but it is not sufficient to eradicate the disease. Policymakers need to also implement other control measures such as case finding, media campaigns, Quarantine and increase in the number of beds in the Ebola treatment centers, good laboratory services, safe burials and social mobilisation, to eradicate the disease.

**Highlights:**

- Due to its high case fatality rate, EVD undoubtedly instills fear in the inhabitants of any affected community.
- We propose an Ebola model with fear, which considers the pathogens in the environment to quantify the effect of fear and environmental transmission on the EVD disease dynamics.
- The fear of death from Ebola is proportional to the Ebola disease transmission rate.
- At high levels of fear, the number of EVD cases decrease.

## 1. Introduction

Ebola virus is a filovirus that causes severe hemorrhagic fever in humans and is believed to be transmitted to humans by animals, Feldmann and Geisbert (2011); Muyembe-Tamfum et al. (2012). Individuals at risk of infection can also contract EVD from the environment by handling or coming into physical contact with pathogen infested objects. Once an individual is infected with the disease, they can develop symptoms after 2 to 21 days of being contaminated, and the infection can last from 4 to 10 days, Astacio et al. (1996). Infected individuals usually have symptoms like headaches, anorexia, lethargy, aching muscles or joints, breathing difficulties, vomiting, diarrhoea, stomach pain, inexplicable bleeding or any sudden inexplicable death, WHO (2014a). Depending on the strength of their immune system, an infected individual can either die immediately or recover after treatment. Even though, research has shown that the virus usually lingers in the semen of some male survivors between about 6-9 months after recovery, Gallagher (2015), recovered individuals become immune to the virus strain they were infected by for at least 10 years, CDC (2015).

Several mathematical modelling studies have been carried out to understand the dynamics and control of EVD, Khan et al. (2015) used a deterministic *SEIHR* (Susceptible–Exposed–Infected–Hospitalised–Recovered) model which differentiates high-risk (e.g. health-workers) and low-risk populations. They estimated the effective contact rate by using an ordinary least-squares estimation to obtain the optimal value of the transmission rate and an estimate of the reproduction number, *R*_0_. In some studies, *SEIR* models were proposed to fit data from the outbreaks in Congo and Uganda, Juga and Nyabadza (2020); Chowell et al. (2004). The model in Chowell et al. (2004) was extended by Legrand et al. (2007), by adding two new compartments for the hospitalized and Ebola deceased individuals who have not yet been buried. In other studies, Althaus (2014); Barbarossa et al. (2015); Lewnard et al. (2014); Djiomba Njankou and Nyabadza (2017), the effect of anti-EVD control measures such as media campaigns, increasing hospitalization, timely burial of people who died from EVD, distribution and use of protective kits in households were incorporated.

Due to its high case fatality rate, EVD undoubtedly instils fear in the inhabitants of any affected community. According to a Bulletin of the WHO in 2016, Van Bortel et al. (2016), infected individuals are psychologically affected and their relatives are traumatized by the infection and the death of the infected individuals. As a result of the fear, inhabitants may become more cautious and resort to preventive measures against the disease such as avoiding physical contact with infected individuals, eating of bush meat, keeping away from public places like schools, hospitals, market places and burial places of the Ebola deceased. These changes in human behaviour resulting from fear of the EVD may lead to a decrease in the human to human, and human to pathogen contact rates. This has significant effects on the disease dynamics and evolution, which the above mentioned mathematical models and other early Ebola models did not consider. Besides, most of them do not take into account the contaminated environment by pathogen infested objects (objects that have come in contact with infected individuals, such as, contaminated syringes used in health care centres, CDC (2014); WHO (2014b,c), bed linen contaminated by infected human’s stool, urine, vomits or sweat of infected individuals, CDC (2014); WHO (2014b)). In this study, we propose an Ebola model with fear, which considers the pathogens in the environment with the aim of quantifying the effects of fear and environmental transmission on the EVD disease dynamics.

The rest of the paper is organised as follows: The introduction is given in Section 1, followed by the model formulation in Section 2. In Section 3, we state and prove the basic properties of the model, find the model steady states and carry out stability analysis of the model steady states. Finally, we carry out numerical simulations in Section 4 and draw relevant conclusions from our results.

## 2. Model formulation

We propose a deterministic model with six independent compartments, namely: susceptible (*S*), infected (*I*), recovered (*R*), deceased (*D*) and a compartment for the pathogens in the environment (*P*). EVD is assumed to be transmitted either by person to person contact or individuals coming in contact with pathogen infested objects (person to pathogen contact). In the event of an Ebola outbreak in a community, the transmission rate is assumed to be reduced by fear that is proportional to the number of deaths. This assumption is driven by the notion that individuals become more cautious as they experience more deaths from EVD within the community. This leads to a decrease in the contact rate. We model the level of fear by the parameter *ϵ*, which measures the impact of fear per dead body. The parameter *ϵ* > 0 with *ϵ* = 0 representing no fear at all, while 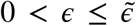 shows increasing levels of fear to a maximum of 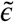. It is plausible to believe that the effective contact rate will reduce with increased levels of fear so that,

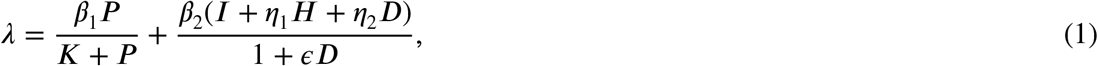

where *K* is the half-saturation constant, that is, the concentration of pathogens that can cause a 50% chance of infection. Here, *β*_1_ and *β*_2_ are the effective contact rates between susceptible individuals and the pathogens in the environment and between susceptible individuals and infected persons that leads to an infection respectively.

The recruitment of susceptible individuals occurs through birth or immigration at a constant rate *π*. Susceptible individuals become infected with the Ebola virus at a rate *λ*. The infectious individuals can either recover at a rate *σ*_1_, or are hospitalised at a rate *σ*_2_ or die at a rate *σ*_3_. We assume that hospitalised individuals are also infectious but with a relatively lower infectivity _1_, compared to that of the deceased and infectious individuals *η*_2_, thus, 0 <*η*_1_ < *η*_2_, with 0 <*η*_1_ < 1. Individuals in the hospitalised class can either recover at a rate *γ*_1_ or die of EVD at a rate *γ*_2_ and the dead bodies of the deceased are safely disposed at a rate *ρ*. Infected individuals and dead bodies of Ebola patients shed pathogens into the environment at rates *α*_1_ and *α*_2_ respectively, while pathogens in the environment decay at a per capita rate *θ*. We assume that people die from natural causes at a rate *µ*. The compartmental diagram for the model is shown in Fig.1. The model derivation is also based on the following additional assumptions:

**Figure 1:**
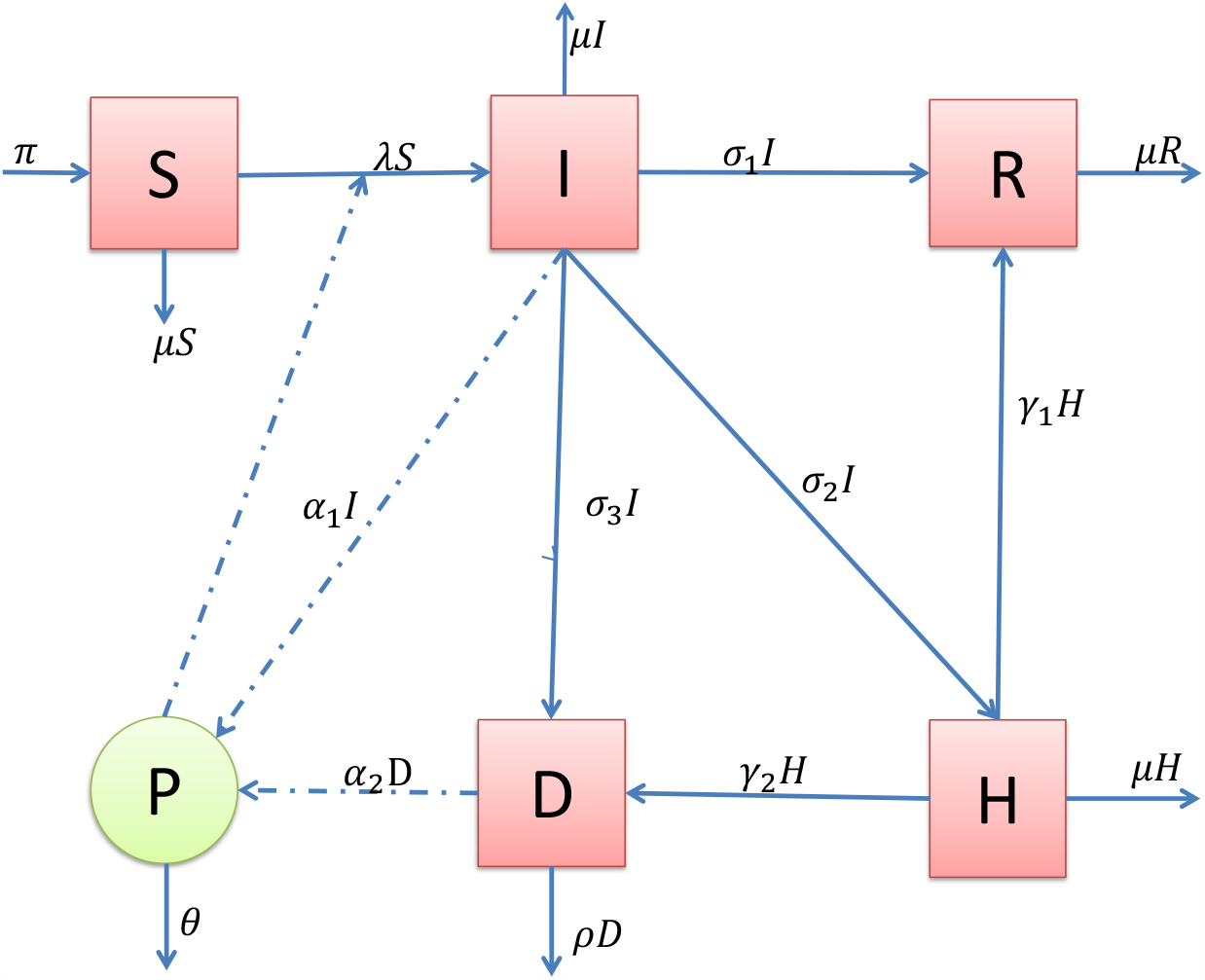
The model diagram for EVD.

The only host population considered in this model is the human population. Animals are not considered as part of the environment, this is justified by the fact that wild animals such as monkeys, apes and duikers, which are also reservoirs for the Ebola virus live in the forests far away from the human habitats, hence there is very little contact between humans and these animals. By definition, the environment is a finite space, thus, it has a carrying capacity. Therefore the increase in the amount of pathogens in the environment as the rate of infection increases is non-linear and saturating, hence the use of a saturating response function in the force of infection. The fear factor does not affect the movement of infectious individuals from the infectious to the hospitalised compartment and from the hospitalised to the dead compartment. This is because naturally, people always take their sick relatives to the hospital irrespective of their condition. Also, a sick person is usually not known to have the Ebola disease until after proper diagnosis in the hospital. The hospitalised individuals are confined in controlled environments and are handled by professionals with protective equipment to handle Ebola patients and dead bodies of Ebola deceased individuals. As such, the level of fear of contracting the disease from the hospitalised individuals is very low and assumed negligible. Also due to their relatively lower infectivity, we assume that the rate at which the hospitalised individuals shed pathogens in the environment is negligible.

The compartmental diagram together with the model assumptions give rise to the following system of differential equations:

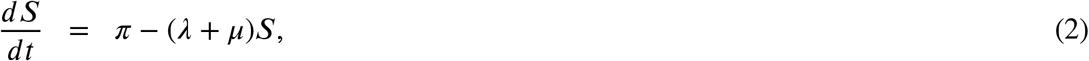

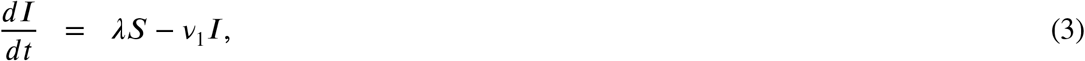

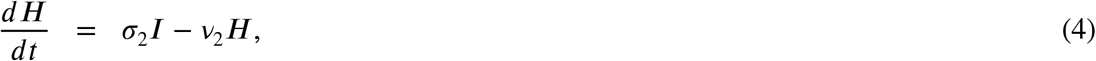

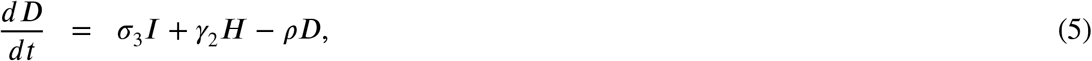

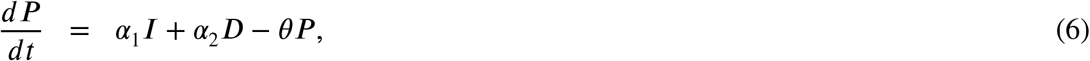

where *v*_1_ = *µ* + *σ*_1_ + *σ*_2_ + *σ*_3_ and *v*_2_ = *µ* +*γ*_1_ +*γ*_2_ with initial conditions, *S*(0) > 0, *I*(0) **≥** 0, *H*(0) **≥** 0, *D*(0) **≥** 0, *P* (0) **≥** 0, for all *t* **≥** 0 and the recovered class is considered to be redundant.

## 3. Properties and analysis of the model

### 3.1. Well posedness

We show that the system is well posed by showing that if the system starts with non-negative initial conditions (*S*_0_, *I*_0_, *H*_0_, *D*_0_, *P*_0_), then the solutions of (2)-(6) will remain non-negative for all *t* ∈ [0, ∞) and that these non-negative solutions are bounded. We thus have the following theorem.

#### Theorem 1.

*Given that S*_0_ > 0, *I*_0_ **≥** 0, *H*_0_ **≥** 0, *D*_0_ **≥** 0, *P*_0_ **≥**0 *and λ* > 0, *the solutions S*(*t*), *I*(*t*), *H*(*t*), *D*(*t*) *and P* (*t*) *of the system (2)-(6) will always be non-negative*.

**Proof 1**. *For the given initial conditions, the first equation gives*

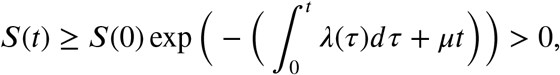

*using the Grownwal inequality. From (3), we have*

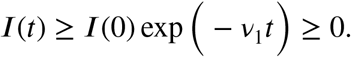

*Similarly*,

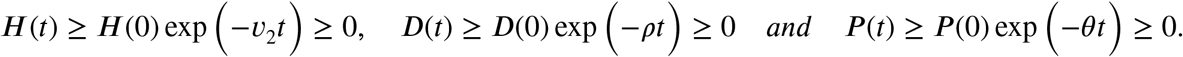

*The solutions of the system (2)-(6) are thus non-negative for any given non-negative initial conditions*.

We now prove the boundedness of the solutions of the system (2)-(6).

#### Theorem 2.

*Suppose m*(*t*) = *S*(*t*) + *I*(*t*) + *H*(*t*) *and the initial conditions for system (2)-(6) satisfy*

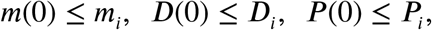

*where*

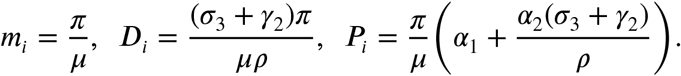

*then the solutions exist and satisfy*

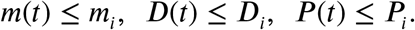

**Proof 2**. *Assume m*(*t*) = *S*(*t*) + *I*(*t*) + *H*(*t*), *then*

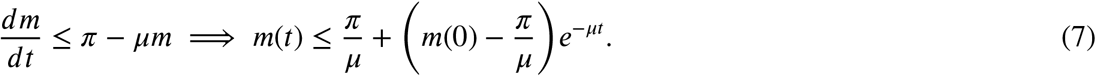

*Thus, m*(*t*) **≤***m*_*i*_. *Substituting the upper bound of m into equation (5) yields*,

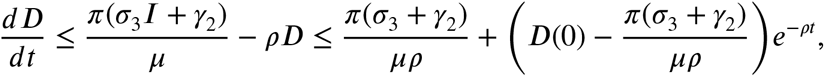

*which implies that D*(*t*) **≤***D*_*i*_. *Similarly*,

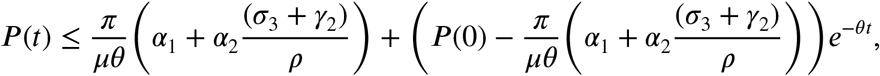

*from which P* (*t*) *P*_*i*_.

Combining the existence and uniqueness of a local solution together with Theorems 1 and 2, we have the following Theorem:

#### Theorem 3.

*The invariant region of the system (2)-(6, where the basic properties of existence, uniqueness and continuity of solutions are valid is the compact set:*

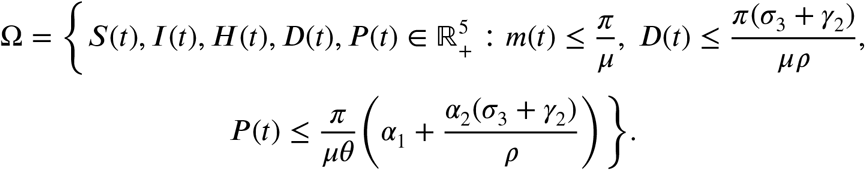

The system is thus well posed and the region *Ω* is therefore a feasible region for our model.

### 3.2. Equilibria of the model

We determine the existence of the equilibrium points of the model (2)-(6) by setting the right hand side of the system (2)-(6) to zero so that

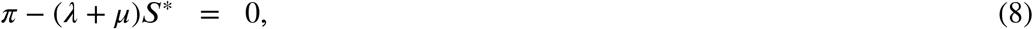

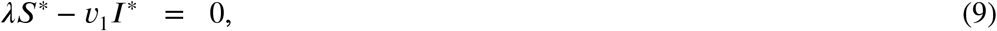

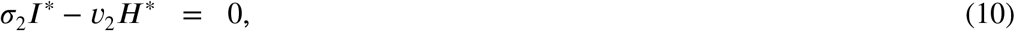

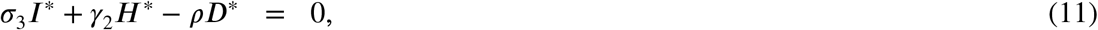

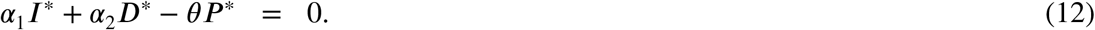

From (10), (11) and (12), we have *H*^*^ = *ϕ*_1_*I*^*^, *D*^*^ = *ϕ*_2_*I*^*^, and *P* ^*^ = *ϕ*_3_*I*^*^, Where 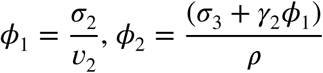, and 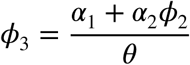.

Substituting the expressions for *H*^*^, *D*^*^, *P* ^*^ into (1), we obtain an expression for *λ* in terms of *I*^*^ so that

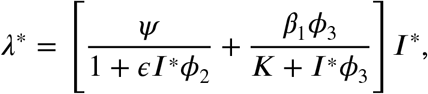

where *Ψ* = *β*_2_(1 +*η*_1_*ϕ*_1_ +*η*_2_*ϕ*_2_. Substituting the expression for *λ*^*^ into (9), we have either

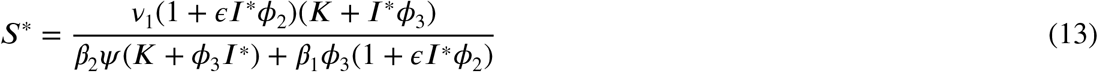

or

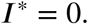

If *I*^*^ = 0, then we have *H*^*^ = *D*^*^ = *P* ^*^ = 0 and 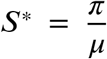. This gives the disease free equilibrium (DFE) point

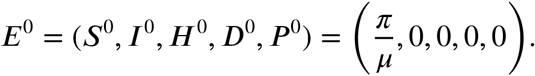

We now determine the reproduction number before determining the endemic steady state using expression (13).

#### 3.2.1. The basic reproduction number (R_0_)

The basic reproduction number denoted by *R*_0_ is defined as the average number of new infections generated by an infected individual or an infected dead body or through contact with a pathogen infested object in a wholly susceptible population, Van den Driessche and Watmough (2002). We use the next generation matrix method to compute *R*_0_ as follows:

By considering the new infections and the transfer matrices (see Van den Driessche and Watmough (2002) for a detailed synopsis), the corresponding Jacobian matrices *F* and *V* evaluated at the DFE are

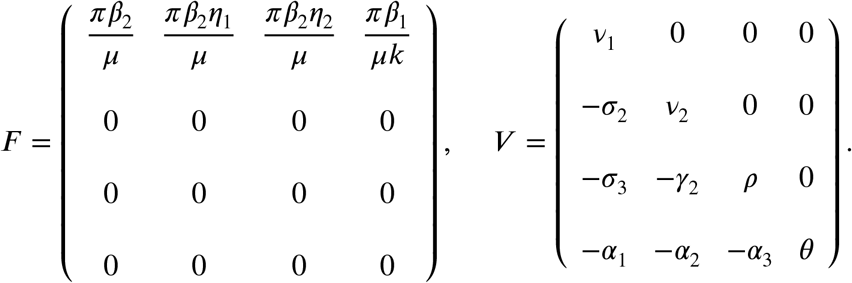

The reproduction number is the spectral radius of the next generation matrix, that is *F V* ^−1^, so that

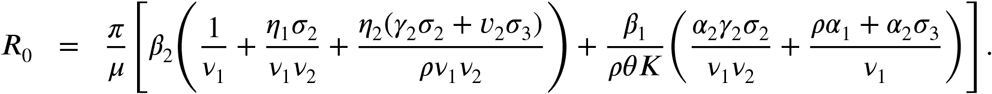

The DFE is also locally stable when *R*_0_ < 1 following Theorem 2 in Mukandavire et al..

#### 3.2.2. The endemic equilibria

Substituting the expression for *S*^*^ in (13) into (8) and simplifying the resulting equation, we obtain the following polynomial.

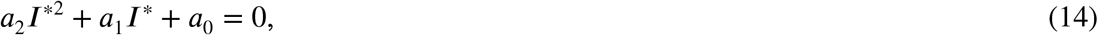

where,

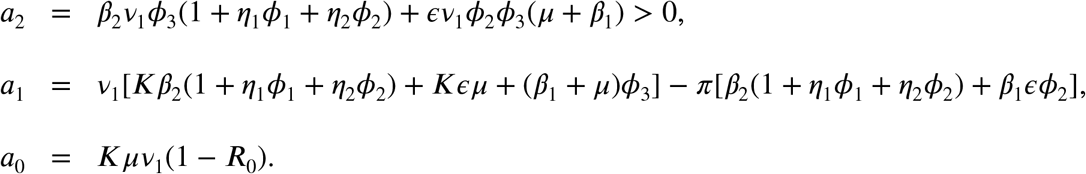

We use Descartes’ law of signs to determine the possible number of positive roots of equation (14) as shown in Table 1.

**Table 1.**
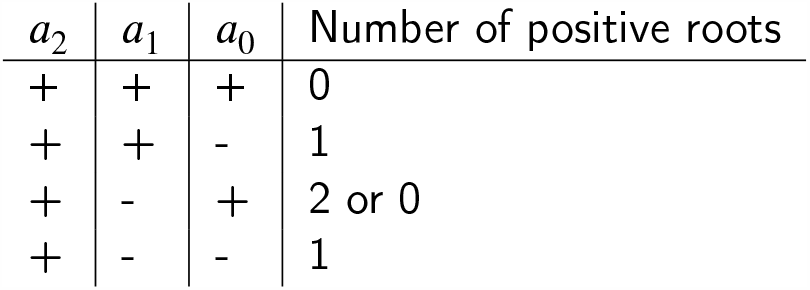
Possible number of positive roots of equation (14)

| $a_2$ | $a_1$ | $a_0$ | Number of positive roots |
| --- | --- | --- | --- |
| + | + | + | 0 |
| + | + | - | 1 |
| + | - | + | 2 or 0 |
| + | - | - | 1 |

The positive solutions of (14) are given by:

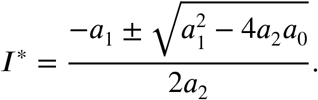

Clearly, the coefficient *a*_2_ is always positive. If *a*_0_ < 0 and 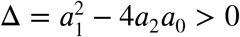, a unique endemic equilibrium exists, irrespective of the sign of *a*_1_. If *a*_0_ > 0, *a*_1_ < 0 and > 0, we get two positive real equilibrium points, otherwise, we have no endemic equilibrium point. The possibility of a backward bifurcation is indicated by the case *a*_0_ > 0, *a*_1_ < 0 and Δ > 0 in which we have two endemic equilibria. We solve the equation Δ = 0 for the threshold value of the reproduction number, 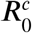, (the value of *R*_0_ below which the DFE is the only steady state) so that

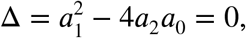

and

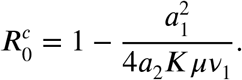

It follows that the system has two positive equilibria for 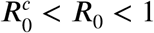.

### 3.3. Bifurcation analysis

#### 3.3.1. The direction of bifurcation

Without loss of generality, assume that *β*_2_ = *cβ*_1_, where *c* is a positive constant. Expressing equation (14) as a function of *β*_1_ and *I*^*^ gives

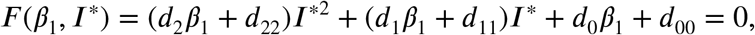

where

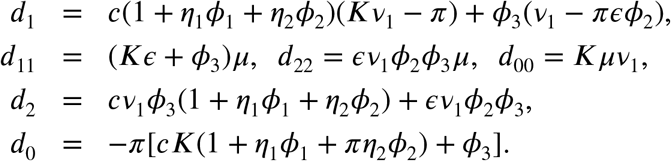

We are interested in the solutions *I*^*^ for a given value of *β*. For *I*^*^ = 0, we get:

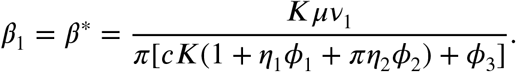

Therefore at *β*_1_ = *β*^*^, *I*^*^ = 0. We compute the bifurcation direction, by definition,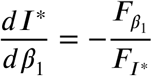, where *Fβ*_1_(*β*^*^, 0) = *d*_*0*_< 0 and *F*_*I*_*(*β*^*^, 0) =*d*_*1*_*β*^*^ + *d*_11_

Note that 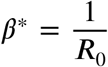. Hence, 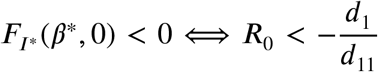 and 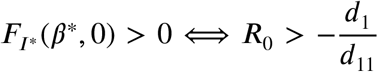. Therefore, if 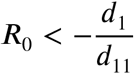, then the bifurcation is backward and if 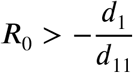, then the bifurcation is forward. We numerically determine the direction of bifurcation in the next section.

#### 3.3.2. Bifurcation simulation

A bifurcation is a change of the topological structure of a system as its parameters pass through a critical value, Kuznetsov (2013). Since the system’s behaviour changes as the reproduction number passes the value 1, *R*_0_ = 1 is a critical point. The occurrence of a backward bifurcation has important public health implications since it might not be enough to reduce *R*_0_ below 1 to eliminate the disease. The basic reproduction number must be further reduced below 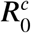 to ensure disease eradication. In this model we have shown that there exists a forward and a backward bifurcation.

Figure 2 (a) and (b) give the graphical representations of the forward and backward bifurcations respectively for the given parameters in the caption. We observe changes in the qualitative behaviour of the system (2)-(6) when *R*_0_ = 1, where *R*_0_ is the bifurcation parameter. For values of *R*_0_ greater than one, we have a forward bifurcation, meaning that the disease will persist in the population and decreasing *R*_0_ to values below one is not a sufficient condition for the disease eradication. The presence of a backward bifurcation makes disease control complicated due to the co-existence of the DFE and the EE for values of *R*_0_ between 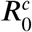 and 1. Thus an increase in the level of fear of death from the EVD is not sufficient to bring about disease eradication. Control measures such as media campaigns, isolation of the infected, and case finding need to be implemented together with the already existing changes in human behaviour due to fear of death, to bring the population to a DFE state. Note that an increase in the level of fear *ε* and a corresponding decrease in the effective contact rate *β*_1_ (with all the other parameters unchanged) changes the backward bifurcation in Figure 2 (b) to a forward bifurcation, as shown in Figure 3.

**Figure 2:**
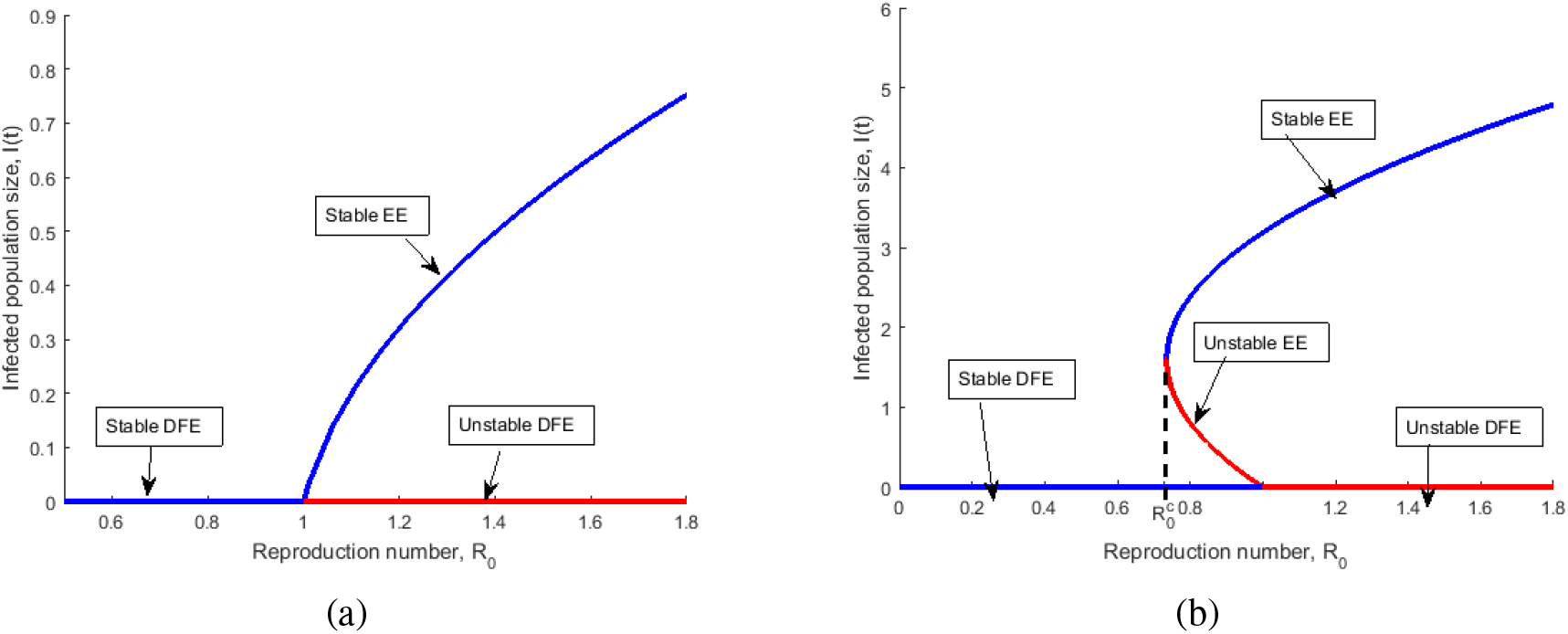
Forward bifurcation in (a) for *ϵ* = 0.95, *β*_1_ = 0.023 and backward bifurcation in (b) for *ϵ* = 0.394, *β*_1_ = 0.25. The rest of the parameter values are the same as the parameter values in Table 3

**Figure 3:**
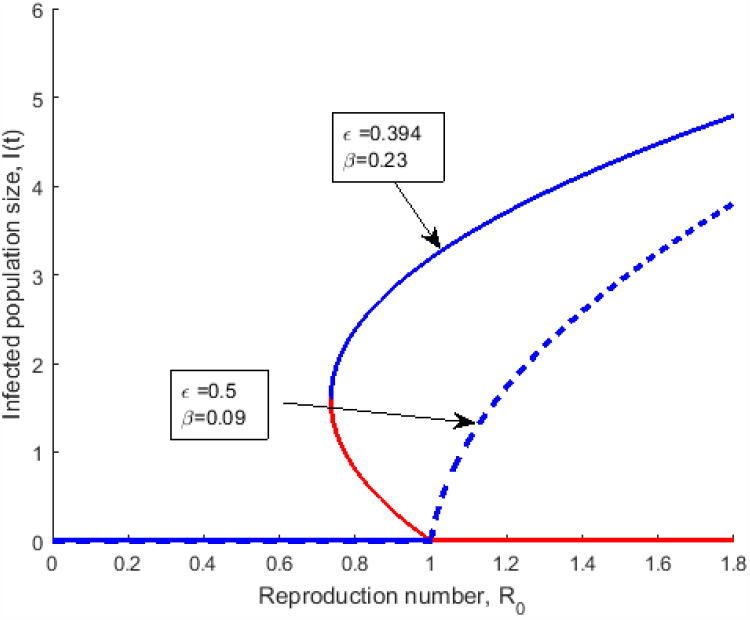
Effect of increasing level of fear, *ϵ*, and decreasing the effective contact rate, *β*_1_, on the infected population size. The other parameters are same as in figure 2.

In Figure 3, the graph with the dashed curve is obtained from Figure 2 (b) by increasing the value of *ϵ* while decreasing the value of *β*_1_. It shows that when more people become afraid of dying, there is a corresponding change in their behaviour, which intend leads to a fall in the effective contact rate. These changes in the values of *ε* and *β*_1_ change a backward bifurcation to a forward bifurcation, thereby making it easier to control and contain the disease.

**Table 3.** Estimated parameter values obtained from the fitting process.

| Parameter | Description | Estimated value (week<sup>-1</sup>) |
| --- | --- | --- |
| $\pi$ | Recruitment rate | 200 |
| $\beta_1$ | Effective human to human contact rate | 0.053 |
| $\beta_2$ | Effective human to pathogen contact rate | 0.064 |
| $\mu$ | Natural death rate | 0.000296 |
| $\sigma_3$ | Disease related death of the infected | 0.6 |
| $\gamma_2$ | Disease related death of the hospitalized | 0.2 |
| $\gamma_1$ | Rate of recovery of the hospitalised | 0.8 |
| $\sigma_1$ | Rate of recovery of the infected | 0.33 |
| $\rho$ | Rate of disposal of dead bodies | 0.009 |
| $\sigma_2$ | Rate of hospitalization of the infectious | 0.019 |
| $\epsilon$ | Level of fear | 0.4 |
| $\theta$ | Rate of removal of pathogens in the environment | 0.016 |
| $K$ | Half saturation constant | 30 |
| $\alpha_1$ | Rate of shedding of pathogens by the infected | 0.05 |
| $\alpha_2$ | Rate if shedding of pathogens by the dead | 0.06 |
| $\eta_1$ | Infectivity rate of the infectious individuals | 0.09 |
| $\eta_2$ | Infectivity rate of the dead individuals | 1.2 |

Many of the epidemiological models that exhibit backward bifurcation have always had the bifur-cation influenced by a single parameter. Our model presents a unique scenario in which backward bifurcation is driven by two important epidemiological parameters. We now present the global stability of the unique endemic equilibrium point in the next subsection.

### 3.4. Global stability of the endemic equilibrium point

#### Theorem 4.

*The endemic equilibrium is globally asymptotically stable in* Ω *if R*_0_ > 1.

**Proof 3**. *If R*_0_ > 1, *we have a unique endemic equilibrium. We consider a candidate Lyapunov function given by*

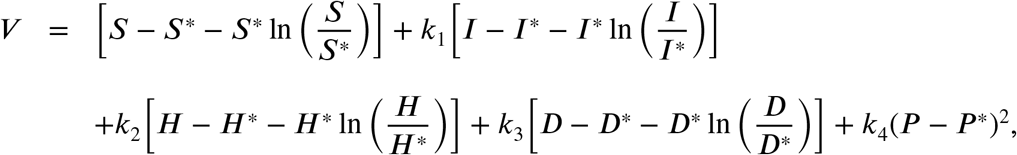

*where k*_1_, *k*_2_, *k*_3_, *k*_4_ *are positive constants to be determined. At endemic equilibrium, the partial derivatives with respect to each variable are*

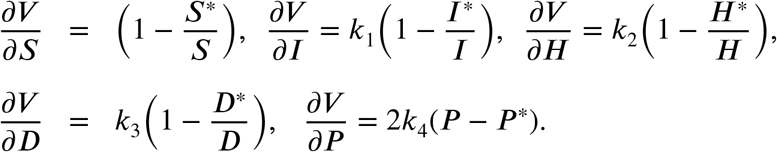

*The endemic equilibrium is clearly a critical point of V*. *The second derivatives are given by*

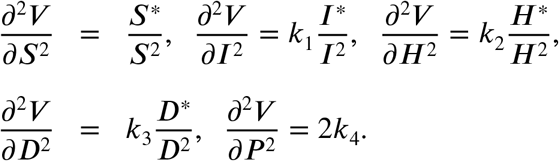

*The second derivatives of V are all positive at any point of* Ω, *therefore, the Lyapunov function V is concave up and the unique endemic equilibrium point is a minimum point of V*. *We now show that* 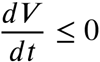. *The time derivative of V is given by*

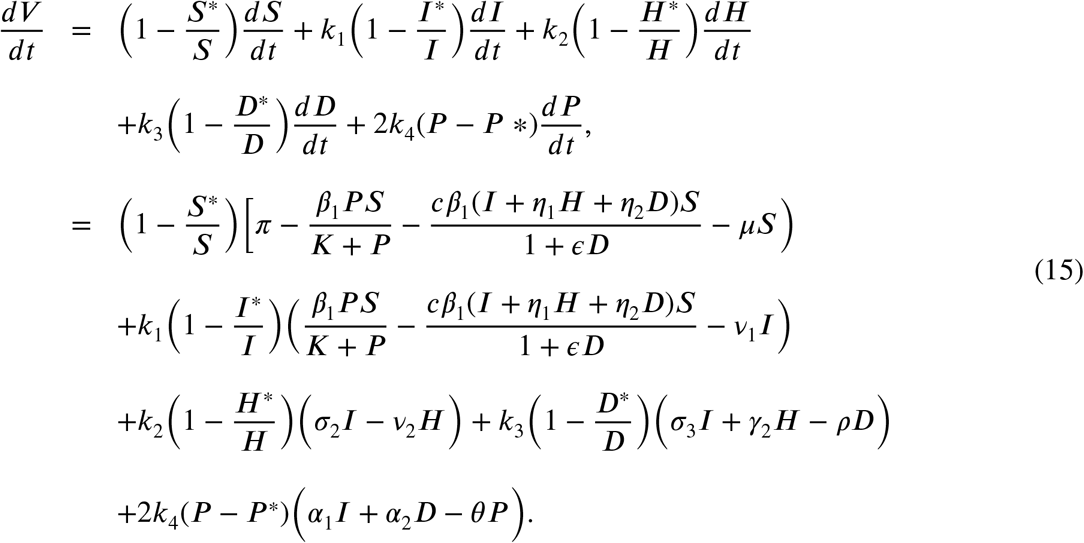

*At the endemic equilibrium, the system (2)-(6) yields the following:*

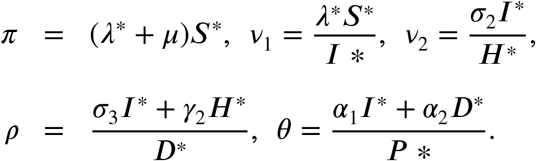

*Substituting the above expressions for the constants π, ρ, θ, v*_1_ *and v*_2_ *into (15), we have*

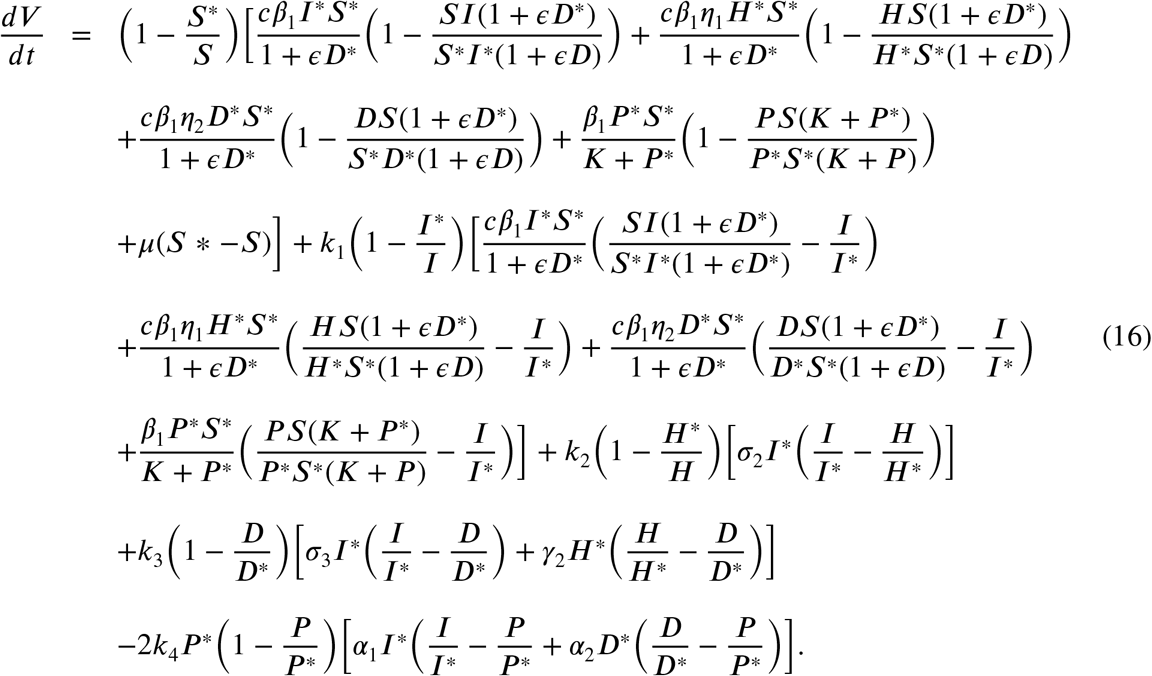

*Let*

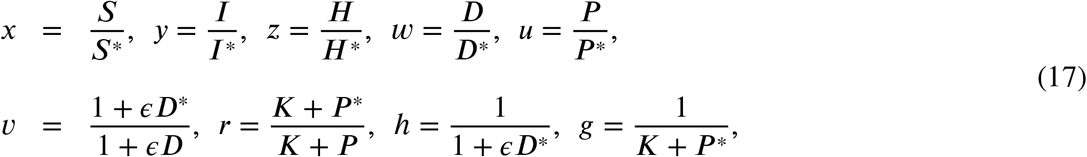

*Substituting the expressions in (17) into (16), we obtain:*

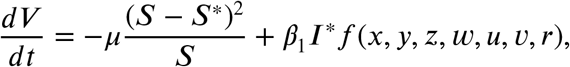

*where*

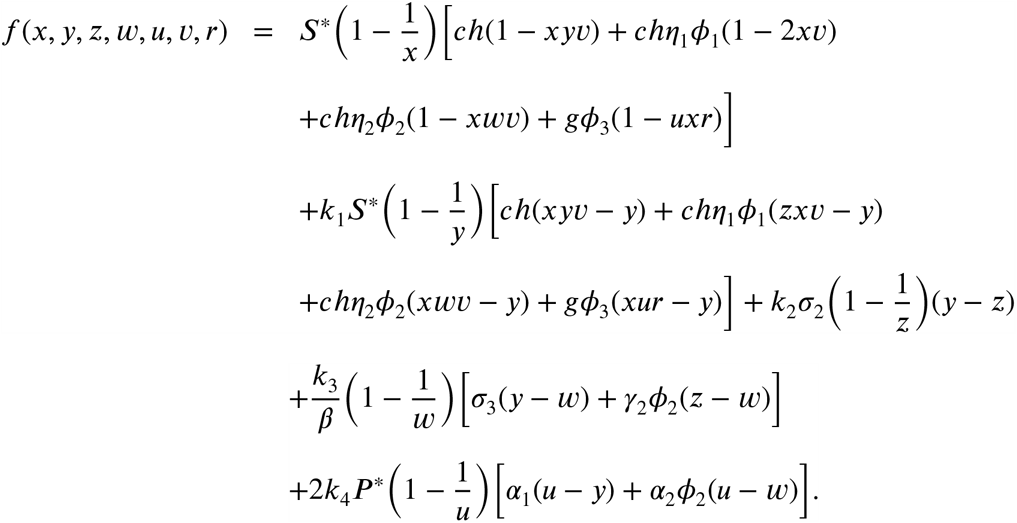

*We equate the coefficients of* 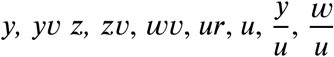, *and w to zero in order to eliminate the positive and non-constant part of f, and solve for the constants k*_1_, *k*_2_, *k*_3_ *and k*_4_ *so that*,

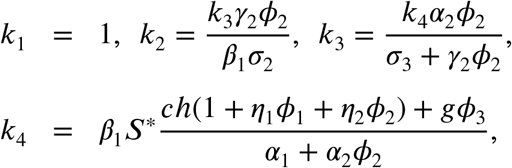

*and*

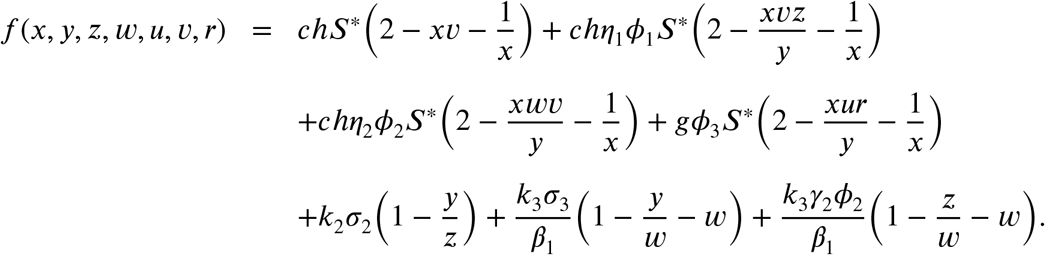

*Given that the expression* 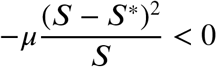, *we show that f* (*x, y, z, w, u,v, r*) < 0.

*Note that the expression* 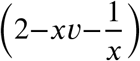 *is less than or equal to zero by the arithmetic-mean-geometric-mean inequality, with equality if and only if x* = = 1. *Also, the expressions* 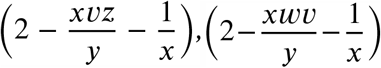, *and* 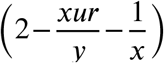 *are less than or equal to zero by the arithmetic-mean-geometric-mean inequality with equality if and only if x* = *y* = *u* = *w* = = *r* = *z* = 1. *After some tedious algebraic manipulations of replacing the constants k*_1_, *k*_2_, *k*_3_ *and k*_4_, *similar conclusions can be drawn for the remaining expressions in f. Therefore f is negative and will be equal to zero if x* = *y* = *z* = *u* = *w* = = *r* = 1. *So V is positive definite at the endemic equilibrium and* 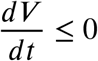 *with equality if and only if S* = *S*^*^, *I* = *I*^*^, *H* = *H* *, *D* = *D*^*^, *P* = *P* ^*^. *The only invariant set contained in Ω is the set containing only the endemic equilibrium point. This shows that each solution which intersects* 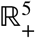 *limits to the endemic equilibrium. Therefore by LaSalle’s invariance principle, LaSalle and Artstein (1976), the endemic equilibrium is globally asymptotically stable on Ω*.

## 4. Numerical simulations

### 4.1. Parameter estimation

In this subsection, we determine the parameter values used in the model. The parameter values are estimated from the fitting process. We fit the model (2)-(6) to the WHO weekly cumulative data from the 2019 and 2020 outbreaks in the North Kivu and South Kivu provinces of the DRC. The data is shown in Table 2. The model presented in the system (2)-(6) is fitted to data in Table 2 using the fminsearch fitting method. Fig 4 shows the optimal fit of the model to the EVD data in Table 2. The sum squares error of the fit is 0.03. Table 3 gives a summary of the parameter values that give the optimal fit.

**Table 2.**
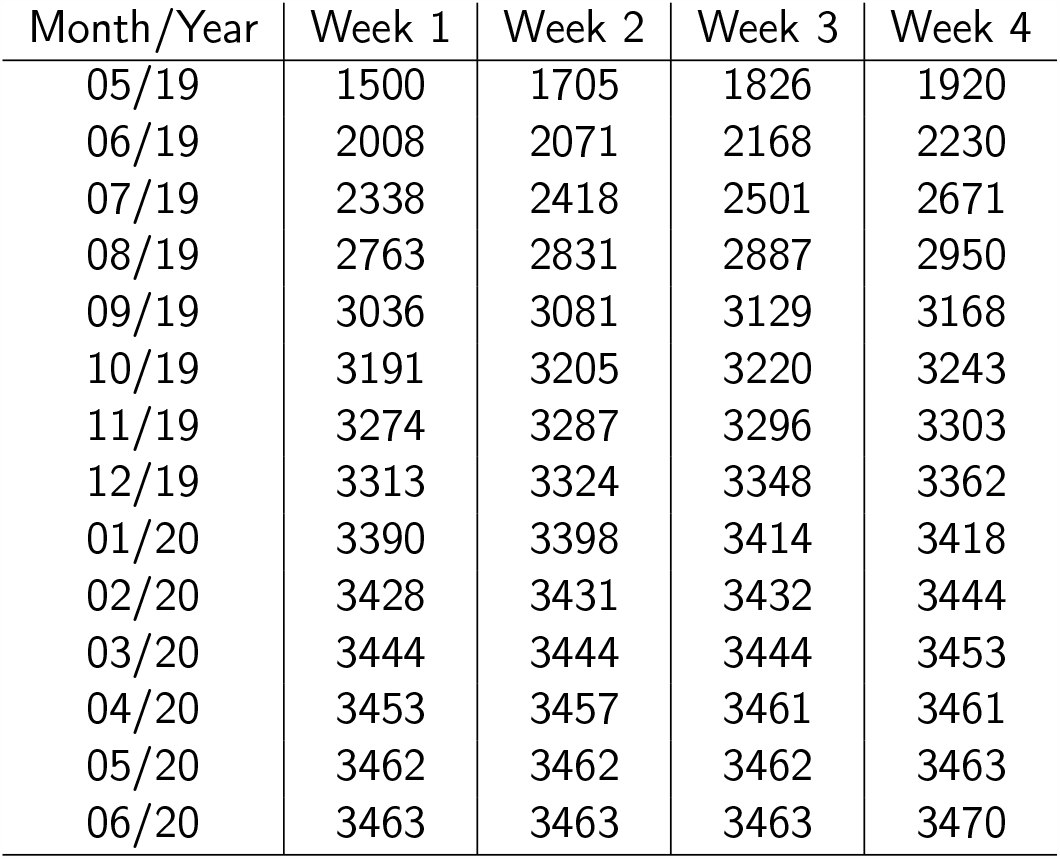
WHO data from the 2019 and 2020 outbreaks in the North Kivu and South Kivu provinces of the DRC.

**Figure 4:**
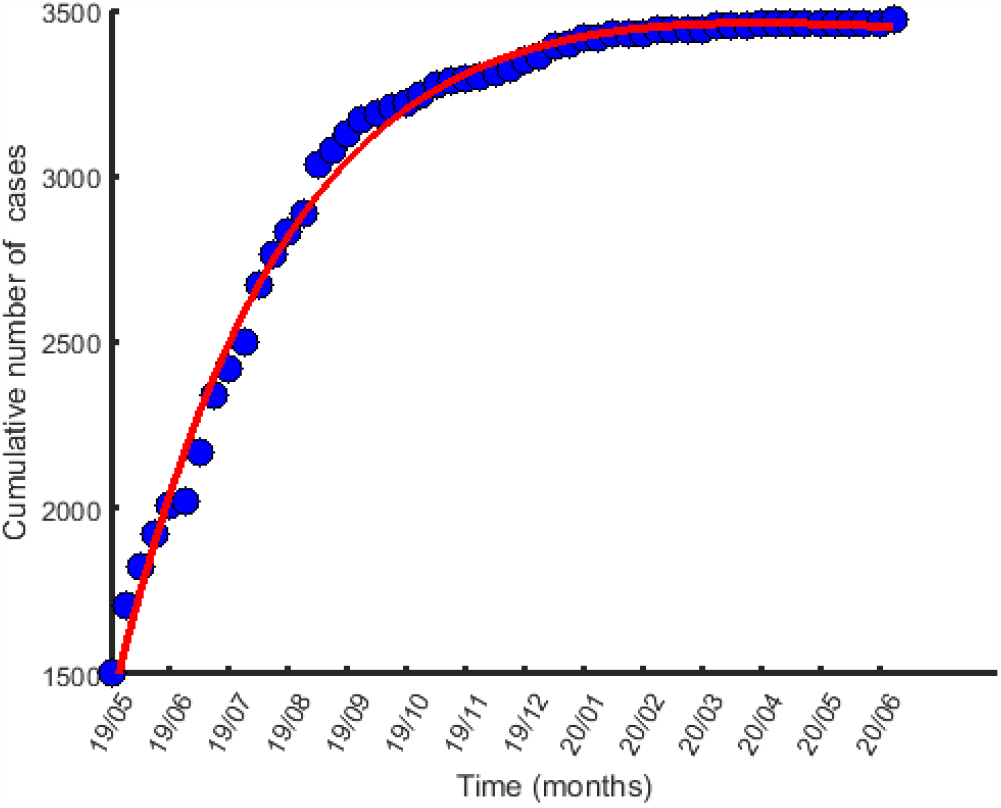
Curve fitting of the model to the data in table 2

### 4.2. Sensitivity analysis

One can not be certain about the parameter values to choose when carrying out numerical simulations of a model. Most of the methods used to collect data from which parameter values of models are chosen are not completely free from errors in as much as a lot of effort is made to minimize errors. These uncertainties are the cause of most of the variability in model predictions. Sensitivity analysis is thus performed to assess this variability in the model predictions, Gomero (2012). We use Latin Hypercube Sampling (LHS) method as described in Blower and othes to explore the entire parameter space of the model. We run the simulations 1000 times to have a large sample size and to increase the level of accuracy of the results. Normally, we are supposed to specify a probability density function (pdf) for each unknown parameter, but since the pdf of each parameter is often unknown, we choose by default a uniform distribution for the variables, independently sample each parameter and do sensitivity plots to determine the most sensitive parameters. The Partial correlation coefficient (PRCC) value for a specific parameter is a Pearson correlation coefficient for the residuals from two regression models. The scatter plots of the most significant parameters are shown in figures 5 and 6.

**Figure 5:**
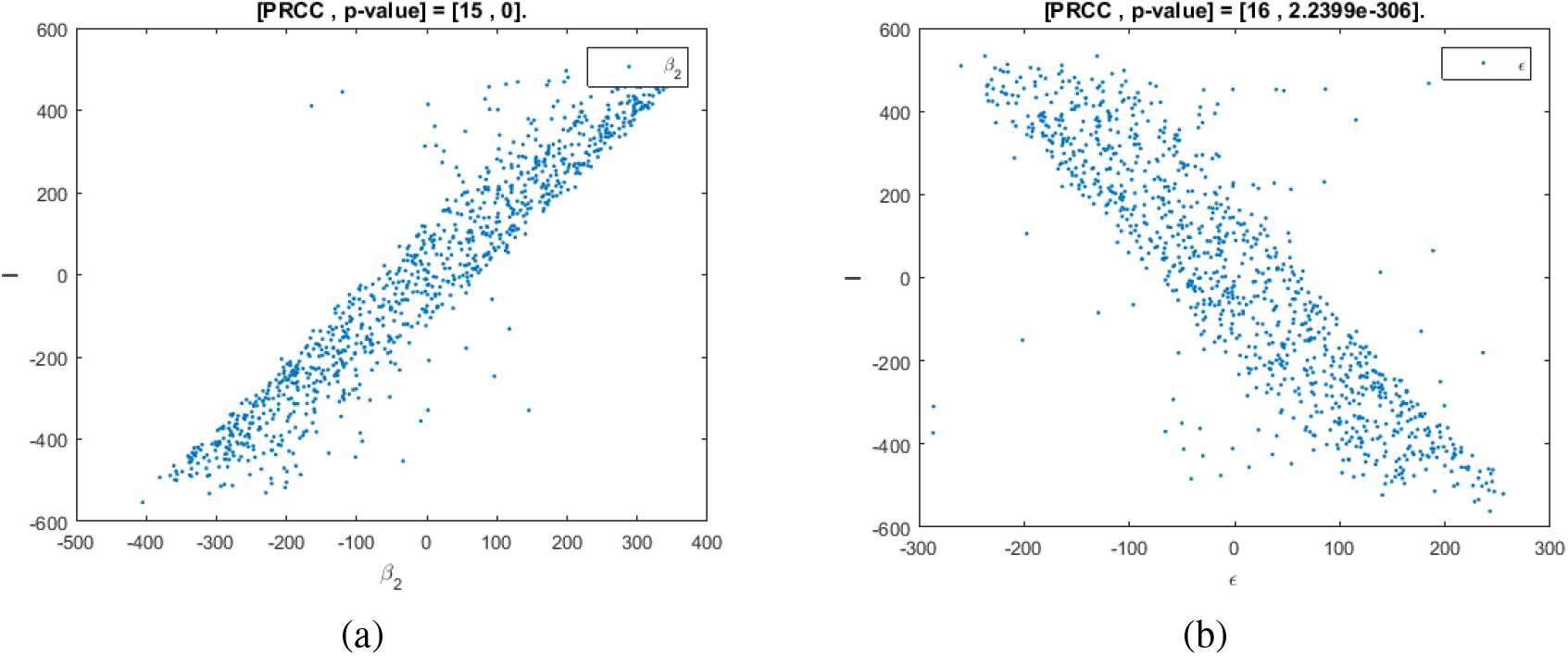
Scatter plots of the infected population *I* as a function of effective person to person contact rate, *β*_2_, in (a), and as a function of the rate of the level of fear in (b)

**Figure 6:**
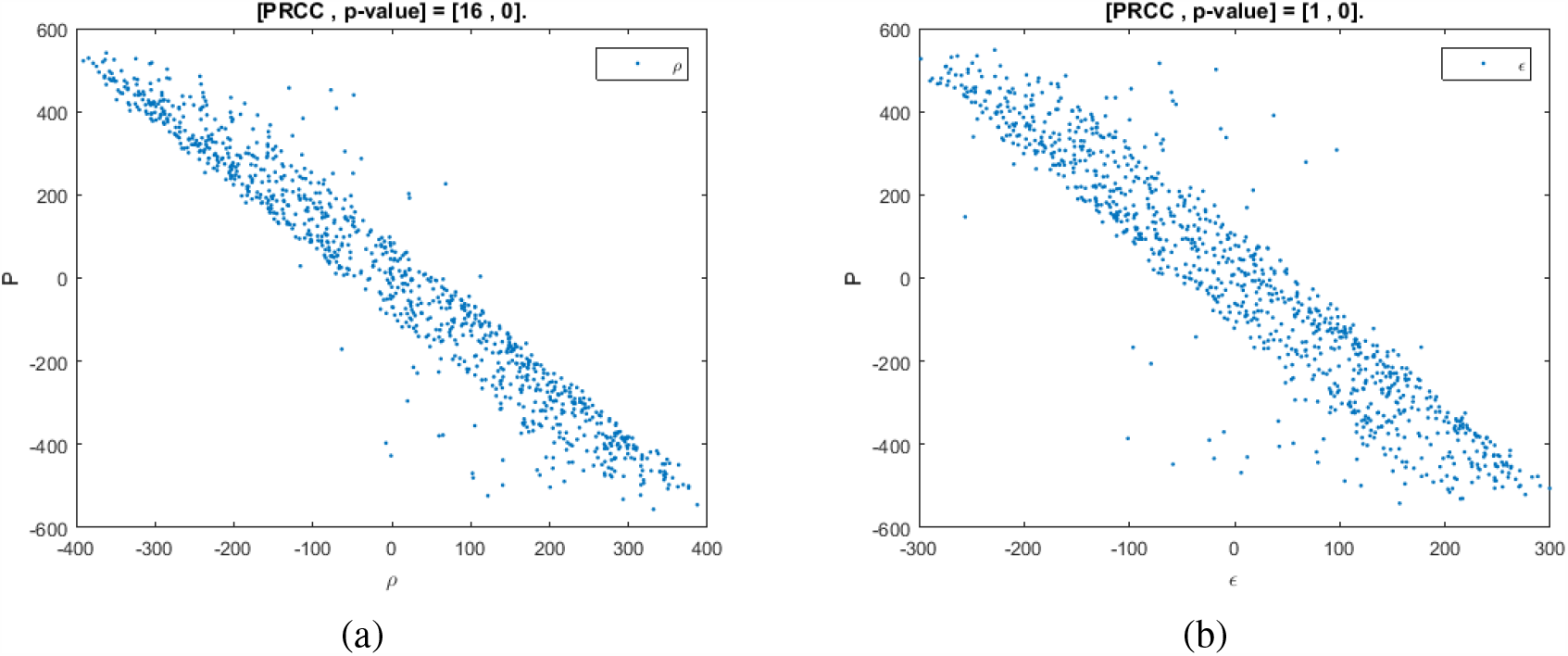
scatter plots of the pathogens in the environment *P* as a function of the rate of save disposal of dead bodies, *ρ*, in (a), and as a function of the rate of the level of fear in (b)

Figure 5 shows the correlations between the most significant parameters and the population of the infected. In Figure 5(a) we obverse an increase in the infected population as the person to person contact rate increases. This is because EVD is easily transmitted when susceptible individuals come in contact with the infectious. In Figure 5(b) we see that the level of fear is negatively correlated to the infected population, however, other control measures such as quarantine, contact tracing and media campaigns are also required to bring about the Ebola disease eradication.

In Figures 6 (a) and (b), we give the scatter plots of the pathogens in the environment *P*, as a function of parameters that are more negatively correlated to *P, ρ* and *ϵ* respectively. The plots show that an increase in the save disposal of dead bodies of Ebola deceased individuals and an increase in the level of fear of dead will help reduce the concentration of pathogens in the environment. Consequently, the infection rate will decrease.

### 4.3. Simulation results

In Figures 7 and 8, we consider the following initial conditions: *S*_0_ = 98000, *I*_0_ = 1500, *H*_0_ = 1000; *D*_0_ = 300; *P*_0_ = 600. The plots are obtained by varying the level of fear, leaving the other parameters constant The observed decrease in the number of infectives and hospitalised as he level of fear increases shows the impact of the fear of death from EVD on the disease transmission dynamics. Also, Figure 9 depicts the impact of fear on the pathogens in the environment. It shows a decrease in the concentration of pathogens in the environment as the level of fear increase. This can be justified by the fact that an increase in the level of fear leads to changes in human behaviour which leads to a decrease in the shedding of pathogens in the environment by the infected humans and infected dead bodies. The contour plot in Figure 10 (a) shows an increase in the reproduction number as the effective contact rates increase. Therefore the disease transmission rate is higher when the contact rates are high. 10 (b) shows the contribution of the pathogens in the environment to disease transmission. We observe that as the infected and the Ebola deceased individuals continually shed pathogens in the environment more people contract the disease. This is seen in the increasing values of *R*_0_ on the *R*_0_-axis.

**Figure 7:**
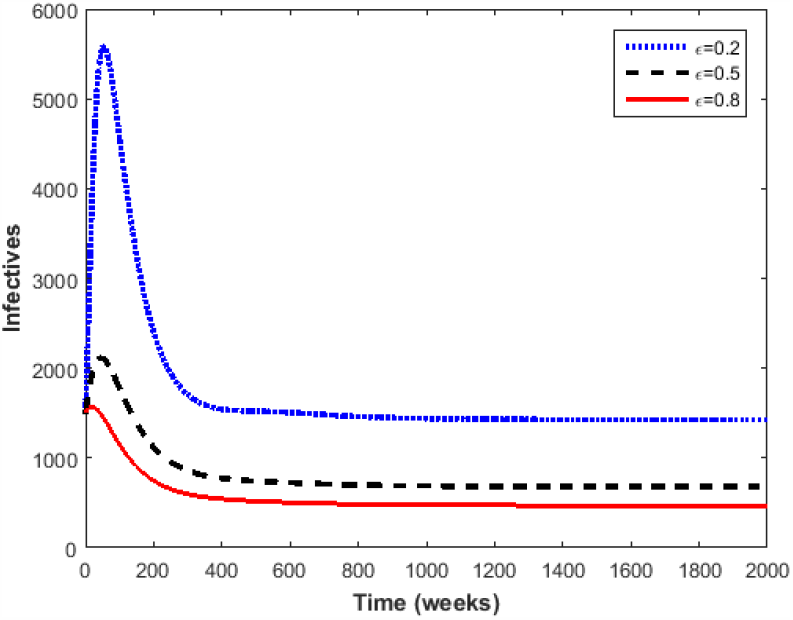
Evolution of the infected population for different levels of fear, *ϵ* The values of the other parameters are the same as the parameter values in Table 3.

**Figure 8:**
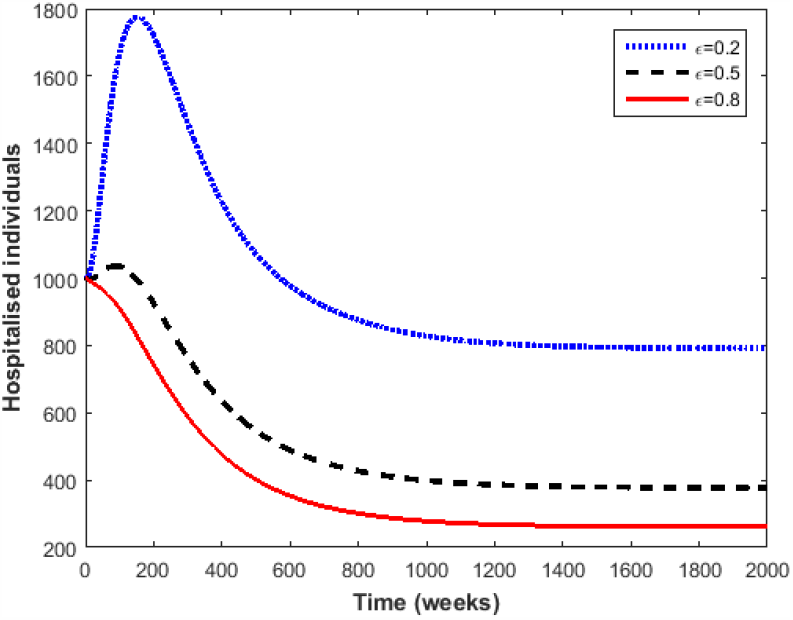
Evolution of the hospitalised population for different levels of fear, *ϵ*. The values of the other parameters are the same as the parameter values in Table 3.

**Figure 9:**
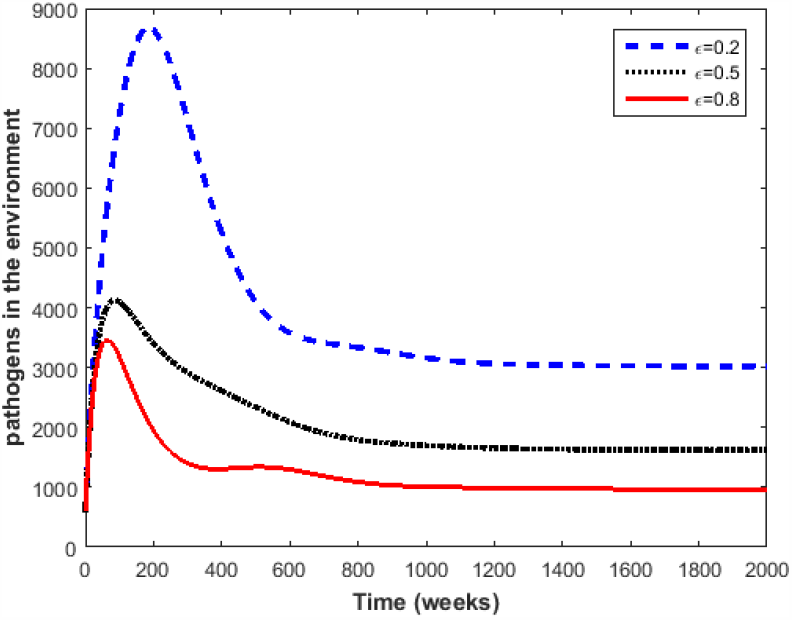
Changes in the concentration of the pathogens in the environment for different levels of fear, *ϵ*. The values of the other parameters are the same as the parameter values in Table 3.

**Figure 10:**
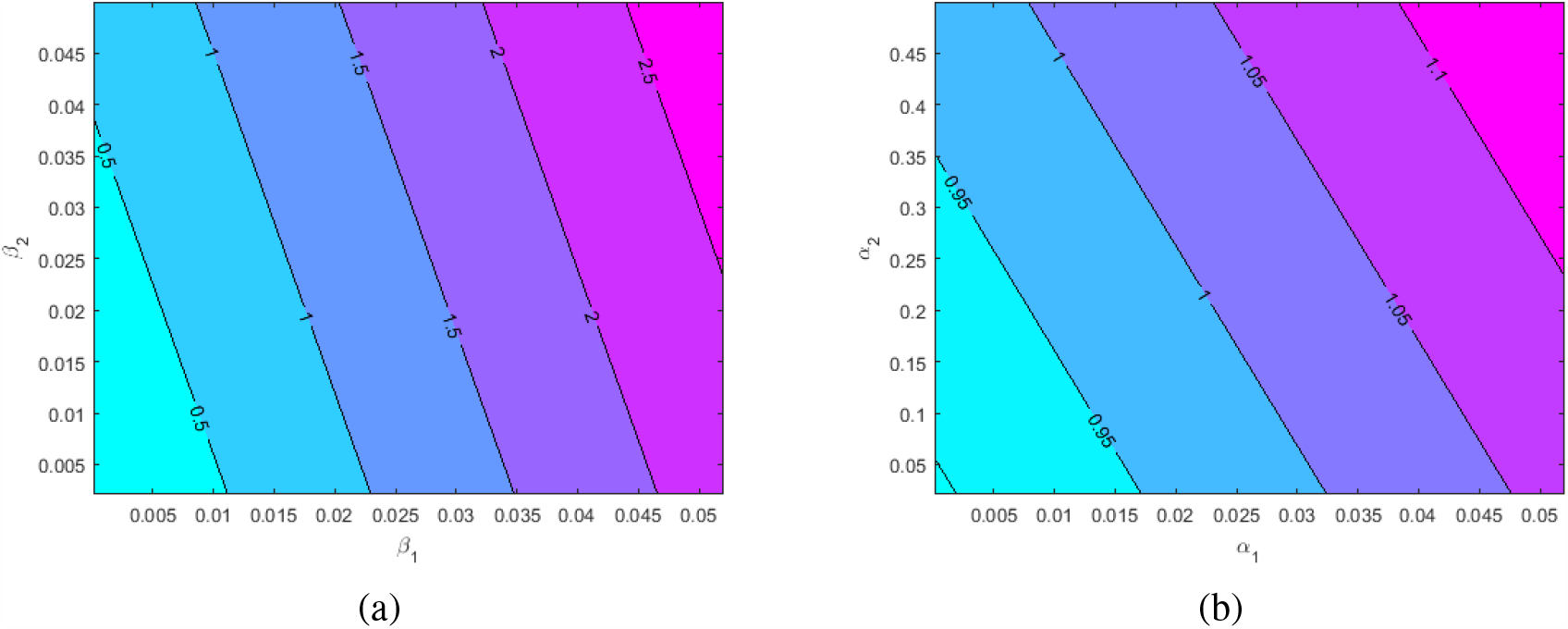
Contour plot of *R*_0_ as a function of effective person to pathogen contact rate, *β*_1_, and effective person to person contact rate *β*_2_ in (a), and as a function of the rate of shedding of pathogens in the environment by the infected and the dead in (b)

## 5. Conclusion

In this work, we presented a 6 compartment deterministic model to assess the impact of fear induced by EVD on the disease transmission dynamics, taking into consideration environmental transmissions. The model’s basic properties such as positivity and boundedness of solutions as well as the reproduction number and steady states were determined and analysed. The analysis of the model reveals the existence of an unstable DFE for values of the reproduction number less than one, or, the existence of a globally stable DFE for values of the reproduction number less than a threshold 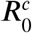, when the level of fear is high. We have also shown that the level of fear in a community experiencing an EVD outbreak is an important factor in disease control. when the level of fear of death from the disease is low, the contact rate increases and controlling EVD becomes more difficult because of the presence of a backward bifurcation. An increase in the level of fear changes human behaviour and leads to a decrease in the effective contact rate, a forward bifurcation appears and reducing the reproduction number to values less than 1 will eradicate EVD. The model fitted well to the DRC (North and South Kivu provinces) cumulative data. One can retain that an increase in the level of fear produces a decrease in the number of EVD cases. However, the impact of fear alone on disease transmission is not significant. It is, therefore, necessary to implement other control measures against EVD like treatment, case-finding and quarantine to limit the disease spread in the population.

The model simulation results show that the level of fear has an inverse relationship with the rate of shedding of pathogens in the environment by the Ebola-infected individuals and dead bodies. This indicates that an increase in the level of fear does not only cause a decrease in the person to person transmission rate but also results to a decrease in the environmental transmission rate (pathogen to person transmission). This model can be improved by considering the growth of the pathogens in the environment independent of the infection dynamics as considered in this manuscript. While the model has some limitations, the consideration of fear and environmental transmission are critical in the management and control of EVD.

## Data Availability

The data used was accessed from
https://www.who.int/emergencies/diseases/ebola/drc-2019

## Declaration

The authors whose names are listed immediately below certify that they have no affiliations with or involvement in any organization or entity with any financial interest (such as honoraria; educational grants; participation in speakers’ bureaus; membership, employment, consultancies, stock ownership, or other equity interest; and expert testimony or patent-licensing arrangements), or non-financial interest (such as personal or professional relationships, affiliations, knowledge or beliefs) in the subject matter or materials discussed in this manuscript.

## Notes

### Competing Interest Statement

The authors have declared no competing interest.

### Clinical Trial

Nil

### Funding Statement

No funding

